# Association of *Streptococcus mutans* harboring bona-fide collagen binding proteins and *Candida albicans* with early childhood caries recurrence

**DOI:** 10.1101/2021.02.28.21252629

**Authors:** B.A. Garcia, N.C. Acosta, S.L. Tomar, L.F.W. Roesch, J.A. Lemos, L.C.F. Mugayar, J. Abranches

**Author notes:** both authors contributed equally (B.A. Garcia, N.C. Acosta). Corresponding author Jacqueline Abranches, Department of Oral Biology, University of Florida College of Dentistry, 1395 Center Drive, PO Box 100424, Gainesville, FL 32610, USA.

## Abstract

Early childhood caries (ECC) recurrence occurs in approximately 40% of treated cases within one year. The association of *Streptococcus mutans* and *Candida albicans* with the onset of ECC is well known. Also, *S. mutans* strains harboring collagen-binding proteins (Cbps) avidly bind to collagen-rich dentin and are linked to increased caries incidence. Here, we investigated the presence of Cbp^+^ *S. mutans* and *C. albicans* in saliva and dental plaque of children with varying caries statuses, as well as the salivary microbiome of these children. In this cross-sectional study, 143 children who were caries-free, treated for ECC with no signs of recurrence after 6 months, or treated for ECC and experiencing recurrence within 6 months following treatment were enrolled. Co-infection with *C. albicans* and *S. mutans*, especially Cbp^+^ *S. mutans,* was strongly associated with caries recurrence. Subjects of the recurrence group infected with Cbp^+^ *S. mutans* showed a greater burden of *C. albicans* and of Mutans streptococci in dentin than those infected with Cbp^−^ *S. mutans*. Microbiome analysis revealed that *Streptococcus parasanguinis* was overrepresented in the caries recurrence group. Our findings indicate that Cbp^+^ *S. mutans* and *C. albicans* are intimately associated with caries recurrence, contributing to the establishment of recalcitrant biofilms.

## INTRODUCTION

Early childhood caries (ECC) is a prevalent chronic disease characterized by the presence of one or more decayed (noncavitated or cavitated lesions), missing (due to caries), or filled tooth surfaces in any primary tooth (dmft) in a child under the age of six^1^ that rapidly and disproportionately affects low- and middle-income countries and communities^2^. Children younger than 3 years of age with any sign of smooth-surface caries, as well children from ages 3 through 5 with dmft score of ≥4 (age 3), ≥5 (age 4), or ≥6 (age 5) surfaces are classified as severe early childhood caries (S-ECC)^1^. The clinical management of ECC often requires surgical interventions, and despite advances in etiology knowledge and treatment approaches, ECC recurrence happens in about 40% of treated cases within one year^3^. This has led to a shift in management from solely operative treatment to more emphasis on prevention and monitoring of caries as a disease^4^.

The dental pathogen *Streptococcus mutans* is consistently associated with caries initiation and development^5^. The cariogenic potential of *S. mutans* resides in three core attributes: (i) the ability to synthesize large quantities of extracellular polymers of glucan from sucrose that aid in the permanent colonization of hard surfaces and in the development of the extracellular polymeric matrix *in situ*, (ii) the ability to transport and metabolize a wide range of carbohydrates into organic acids (acidogenicity), and (iii) the ability to thrive under environmental stress conditions, particularly low pH (aciduricity)^6^. Strains of *S. mutans* are classified into four serotypes (*c, e, f* and *k*) with over 70% of strains isolated from dental plaque belonging to serotype *c*, 20% to serotype *e*, and the remaining to serotyped *f* and *k*^6^. Most individuals usually harbor a single serotype of *S. mutans*, however colonization with multiple serotypes has been reported^7^. While the overall detection of *S. mutans* in saliva is not a strong predictor of caries risk^8^, it has been proposed that the presence of *S. mutans* harboring bona-fide collagen-binding proteins (Cbps) Cnm or Cbm can be linked to increased caries risk^9^. Specifically, Cbp^+^ *S. mutans* is associated with increased caries incidence and poor caries outcomes in children^9–11^. While Cnm and Cbm are found in nearly 20% of *S. mutans* isolates^10^, they are thought to be prevalent in the less common serotypes *e, f* and *k* (~85% isolates)^10^. Importantly, the presence of Cbps was associated with the expression of virulence traits that facilitate the persistence of *S. mutans* in the oral cavity such as adhesion to and invasion of oral cell lines and binding to collagen-rich surfaces (dentin and roots), that ultimately resulted in increased caries severity in a rat caries model^11^.

Clinical studies have also shown a strong association of *Candida albicans* with ECC^12–14^. In fact, Xiao *et al*.^14^, reported that children colonized with *C. albicans* have approximately 5 times greater risk of developing ECC than those that are not infected^14^. Moreover, it is well known that co-infection with *S. mutans* and *C. albicans* is strongly associated with severe caries in children (S-ECC)^14–17^. These clinical observations are supported by an *in vivo* study showing that co-infection with *S. mutans* and *C. albicans* can cause more severe and extensive caries in rats^18^. Thus, a better understanding of the dynamics of cariogenic oral biofilm in patients experiencing ECC is paramount to identify optimal methods for prevention of caries initiation, progression and recurrence.

The present study builds on the premise that the association of *S. mutans* and *C. albicans* can result in hypervirulent biofilms that are intimately associated ECC; and that Cbp^+^ *S. mutans* can colonize multiple sites in the oral cavity, especially collagen-rich surfaces such as dentin, contributing to *S. mutans* niche expansion and caries severity. Here, to determine whether co-infection with *S. mutans* and *C. albicans* is also associated with ECC recurrence, we investigated the presence of *S. mutans,* including Cbp^+^ strains, and *C. albicans* in saliva and dental plaque of children who were caries free (CF), caries experienced without recurrence (CE-NR), and had active recurrent caries (CR). In addition, to gain insight on the microbiota associated with ECC recurrence and in an attempt to find potential biomarkers for caries recurrence, the salivary microbiome of children with different caries statuses was also investigated.

## RESULTS

### Study Sample Characteristics

A total of 143 children [73 caries free (CF), 45 caries experienced without recurrence (CE-NR) and 25 with active recurrent caries (CR)] were enrolled in the study (Table 1). The age average of all participants was 40 months, but children in the CE-NR group were significantly older (average of 47.9 months) than the CF and CR groups (p<.0001). Then, because of the age difference between CE-NR and the other two groups, age in months was included as a covariate in the subsequent analyses. There was no statistically significant difference in the biological sex distribution among the three groups [X^2^ (2, N=143) =0.01, p=.99]. About 38% of the children enrolled were non-Hispanic White, 23% were non-Hispanic Black or African American, and 14% were Hispanic. CF group had a significant greater proportion of non-Hispanic Black or African American children (45.21%), while the CE-NR and CR groups were composed of a significantly larger proportion of Hispanic children (24.44%) and other or not reported race/ethnicities (56%), respectively [X^2^ (6, N=143)=61.54, p<.0001]. Of the 70 children with ECC in the CE-NR and CR groups, most of them (82.85%) had severe ECC (S-ECC), with dmft averages of 8.9 and 8.7, respectively (Table 1).

**Table 1.** Selected demographic and clinical characteristics of the study subjects.

| Variable | Groups |  |  |  | All subjects |
| --- | --- | --- | --- | --- | --- |
|  | CF (n = 73) | CE-NR (n = 45) | CR (n = 25) | P value | (n = 143) |
| Age in months, mean ± SD | 35.8 ± 16.2 | 47.8 ± 9.1 | 38.5 ± 12.7 | <.0001<sup>a</sup> | 40.0 ± 14.7 |
| Sex |  |  |  | .99<sup>b</sup> |  |
| Female | 37 (50.7) | 23 (51.1) | 13 (52.0) |  | 73 (51.0) |
| Male | 36 (49.3) | 22 (48.9) | 12 (48.0) |  | 70 (49.0) |
| Race/ethnicity |  |  |  | <.0001<sup>b</sup> |  |
| White, non-Hispanic | 30 (41.1) | 17 (37.8) | 8 (32.0) |  | 55 (38.4) |
| Black or African American, non-Hispanic | 33 (45.2) | 0 (0.0) | 0 (0.0) |  | 33 (23.1) |
| Hispanic | 6 (8.2) | 11 (24.44) | 3 (12.0) |  | 20 (13.9) |
| Other or not reported | 4 (5.5) | 17 (37.8) | 14 (56.0) |  | 35 (24.4) |
| dmft | 0 ± 0 | 8.9 ± 5.0 | 8.7 ± 5.2 | <.0001<sup>a,c</sup> | 4.3 ± 5.6 |
| dmft / erupted teeth, % ± SD | 0 ± 0 | 50.9 ± 30.7 | 45.9 ± 26.6 | <.0001<sup>a,c</sup> | 24.0 ± 32.0 |
| S-ECC | 0 (0.0) | 36 (80.0) | 22 (88.0) | <.0001<sup>b,c</sup> | 58 (40.6) |

Values are presented as mean ± SD or number (%).
CF = caries free; CE-NR = caries experienced - no recurrence; CR = active caries- recurrence; dmft = decayed, missing (due to caries), or and filled primary teeth; S-ECC = severe early childhood caries.
<sup>a</sup>Generalized linear modeling
<sup>b</sup>Chi-square test
<sup>c</sup>Adjusted for age

### Microbiological analysis reveals that CR subjects have an increased burden of Mutans streptococci and *Candida* spp. in plaque

To estimate the infection levels with Mutans streptococci and *Candida* spp. among the three groups, we collected saliva and plaque samples from enamel surfaces from all children enrolled in this study. No statistically significant differences in Mutans streptococci and *Candida* spp. counts were found among the three groups in saliva. However, the CR group showed higher counts of Mutans streptococci and *Candida* spp. in healthy plaque (H) than the CF subjects (p<.0001) (Fig. 1a–b). Additionally, to determine the infection burden in diseased sites, we collected plaque samples from white spot lesions (WS) on the edge of a carious lesion and from cavitated lesions that visibly extend into the dentin (D) from CR children and found that dentin plaque presented the highest counts of Mutans streptococci and *Candida* spp. when compared to white spot and healthy plaques (p<.05) (Fig. 1b).

**Figure 1.**
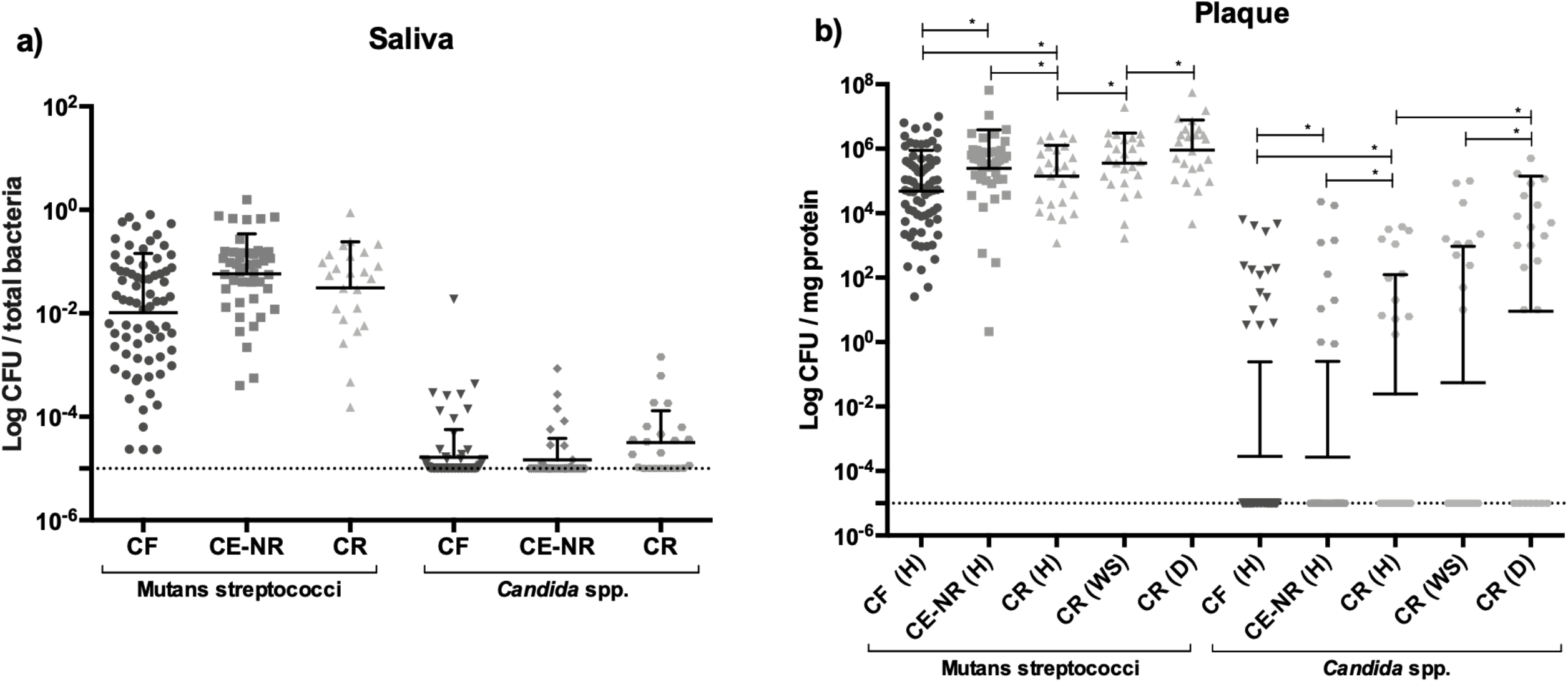
Microbiological analysis of saliva and plaque samples from caries free (CF), caries experienced - no recurrence (CE-NR), and active recurrent caries (CR) groups. Panels A and B depict Mutans streptococci and *Candida* spp. counts in saliva samples and in healthy plaque (H), white spot lesion plaque (WS) and cavitated dentin plaque (D) from the study groups, respectively. The CFU counts among the three caries status groups were compared using general linear mixed modeling, controlling by age. Data shown are mean ± SD values of log total colony count of Mutans streptococci and *Candida* spp. colonies.

### Presence of Cbp^+^ *S. mutans* is strongly associated with ECC relapse

Next, we determined the prevalence of *S. mutans*, including strains harboring genes coding for *cnm* or *cbm*, and its serotypes in the study groups. When compared to CF and CE-NR groups, children in the CR group had a larger prevalence of *S. mutans* (80%) [X^2^(2, N=143)=99.84, p<.0001] (Table 2). All *S. mutans* isolates were classified as serotypes *c, e, f* or *k* with serotype *c* being the most prevalent in this cohort (20.3% of all children). Although the presence of multiple serotypes (*c* + *e* and *e* + *k*) was found in some children (7.7% of all subjects), most of the them were infected with a single serotype. Infection with serotype *c* [X^2^(2, N=143)=31.13, p<.0001] or mixed infection with serotypes *e* and *k* [X^2^(2, N=143)=9.03, p=.01] were associated with caries relapse (Table 2). Strikingly, *cnm*^+^ or *cbm*^+^ (Cbp^+^) *S. mutans* strains were only found in CR subjects and were strongly associated with the ECC recurrence group [X^2^(2, N=143)=28.23, p<.0001] (Table 2). Of a total of 25 CR cases, *cnm*^+^ strains were found in 20% of the subjects and *cbm*^*+*^ strains in 8% of the subjects; most of the isolates harboring the Cbp genes (*cnm* or *cbm*) belonged to serotype *c* (71.4%). The overall prevalence of Cbp^+^ strains (*cnm*^+^ or *cbm*^+^) among the *S. mutans* strains obtained in our study was about 15% (Table 2). When breaking down the CR group into subjects infected with Cbp^+^ and Cbp^−^ *S. mutans* and comparing their demographics and clinical characteristics as depicted in table 3, no significant differences regarding age, sex, dmft score and severity of ECC were found (p>.05). Finally, all children infected with Cbp^*+*^ *S. mutans* strains (*cnm*^*+*^ or *cbm*^*+*^) were younger than 3 years of age (mean age ± SD = 27.8 ± 5 months) and classified as S-ECC cases (Table 3).

**Table 2.** Presence of *S. mutans*, its molecular characterization, and presence of *Candida* spp.

| Variable | Groups |  |  |  | All subjects |
| --- | --- | --- | --- | --- | --- |
|  | CF (n = 73) | CE-NR (n = 45) | CR (n = 25) | P value* | (n = 143) |
|  | n (%) | n (%) | n (%) |  |  |
| *S. mutans* presence | 11 (15.1) | 14 (31.1) | 20 (80.0) | <.0001 | 45 (31.5) |
| Serotype distribution |  |  |  |  |  |
| Ser c only | 6 (8.2) | 8 (17.8) | 15 (60.0) | <.0001 | 29 (20.3) |
| Ser e only | 1 (1.4) | 2 (4.4) | 1 (4.0) | .56 | 4 (2.8) |
| Ser f only | 0 (0.0) | 1 (2.2) | 0 (0.0) | .33 | 1 (0.7) |
| Ser k only | 0 (0.0) | 0 (0.0) | 0 (0.0) |  | 0 (0.0) |
| Ser c + e | 1 (1.4) | 3 (6.7) | 0 (0.0) | .15 | 4 (2.8) |
| Ser e + k | 3 (4.1) | 0 (0.0) | 4 (16.0) | .01 | 7 (4.9) |
| Genes coding for Collagen Binding Proteins |  |  |  |  |  |
| Cbp (*cnm* + *cbm*) | 0 (0.0) | 0 (0.0) | 7 (28.0) | <.0001 | 7 (4.9) |
| *cnm* | 0 (0.0) | 0 (0.0) | 5 (20.0) | .0002 | 6 (4.2) |
| *cbm* | 0 (0.0) | 0 (0.0) | 2 (8.0) | .03 | 2 (1.4) |
| *Candida* spp. presence | 29 (39.7) | 16 (35.6) | 20 (80.0) | .0007 | 65 (45.5) |
| *Candida albicans* presence | 15 (20.6) | 9 (20.0) | 17 (68.0) | <.0001 | 41 (28.7) |
| Presence of more than one species of *Candida*. | 9 (12.3) | 2 (4.4) | 5 (20.0) | .30 | 16 (11.2) |
| Co-infection with *S. mutans* and *C. albicans* | 7 (9.6) | 7 (15.6) | 17 (68.0) | <.0001 | 31 (21.7) |

CF = caries free; CE-NR = caries experienced - no recurrence; CR = active caries - recurrence
\*P-values based on Cochran-Mantel-Haenszel test of general association, controlling for age by stratification

**Table 3.** Demographic and clinical characteristics of the subjects with caries recurrence.

| Variable | Caries recurrence group |  |  | Total subjects CR group (n = 25) |
| --- | --- | --- | --- | --- |
|  | CBP<sup>-</sup> (n = 18) | CBP<sup>+</sup> (n = 7) | P value |  |
| Age in months, mean ± SD | 42 ± 13 | 27.8 ± 5 | .12 | 38.5 ± 12.7 |
| Sex |  |  | .19 |  |
| Female | 8 (44.4) | 5 (71.4) |  | 13 (52.0) |
| Male | 10 (55.6) | 2 (28.6) |  | 12 (48.0) |
| Race/ethnicity |  |  | .03 |  |
| White, non-Hispanic | 8 (44.4) | 0 (0.0) |  | 8 (32.0) |
| Black or African American, non-Hispanic | 0 (0.0) | 0 (0.0) |  | 0 (0.0) |
| Hispanic | 1 (5.6) | 2 (28.6) |  | 3 (12.0) |
| Other or not reported | 9 (50.0) | 5 (71.4) |  | 14 (56.0) |
| dmft | 9.3 ± 5.7 | 7.1 ± 3.4 | .45 | 8.7 ± 5.2 |
| dmft / erupted teeth, % ± SD | 49.1 ± 29.0 | 37.7 ± 18.2 | .40 | 45.9 ± 26.6 |
| S-ECC | 15 (83.3) | 7 (100.0) | .59 | 22 (88.0) |

Values are presented as mean ± SD or number (%).
CF = caries free; CE-NR = caries experienced - no recurrence; CR = active caries – recurrence; dmft = decayed, missing (due to caries), or and filled primary teeth; S-ECC = severe early childhood caries.

### Co-infection with *C. albicans* and *S. mutans* is strongly associated with caries relapse

We evaluated the prevalence of infection with *Candida* spp. among the different study groups. About 80% of the subjects in the CR group were infected with *Candida* spp., an incidence that was significantly greater than that observed for the CF and CE-NR groups in which *Candida* spp. were found in less than 40% of the subjects [X^2^(2, N=143)=14.49, p=.0007] (Table 2). At species level, *C. albicans* was isolated in 68% of the CR subjects whereas it was found in only 20% of the CF and CE-NR subjects [X^2^(2, N=143)=21.41, p<.0001]. Importantly, table 2 shows that co-infection with *S. mutans* and *C. albicans* was most prevalent in the CR group compared to CF and CE-NR [X^2^(2, N=143)=39.42, p<.0001].

### CR children infected with Cbp^+^ *S. mutans* strains have a higher burden of Mutans streptococci and *Candida* spp. than those infected with Cbp^−^ strains

Since we observed that Cbp^+^ *S. mutans* strains were only found in the CR group, we compared the infection characteristics of CR subjects infected with Cbp^+^ *S. mutans* versus those infected with Cbp^−^ *S. mutans*. We observed that subjects infected with Cbp^+^ *S. mutans* tended to have overall higher counts of *Candida* spp. and Mutans streptococci in white spot lesions (WS) or healthy plaque (H) than those who were colonized with Cbp^−^ *S. mutans* (Fig. 2), but the differences were not statistically significant. However, in dentin sites, Mutans streptococci counts were significantly higher (8-fold) in subjects infected with Cbp^+^ *S. mutans* in comparison to subjects that were infected Cbp^−^ *S. mutans* (p=.0077). Although not statistically significant, *Candida* spp. counts were 13-fold higher in dentin in the Cbp^+^ *S. mutans* group than in Cbp^−^ *S. mutans* subjects.

**Figure 2.**
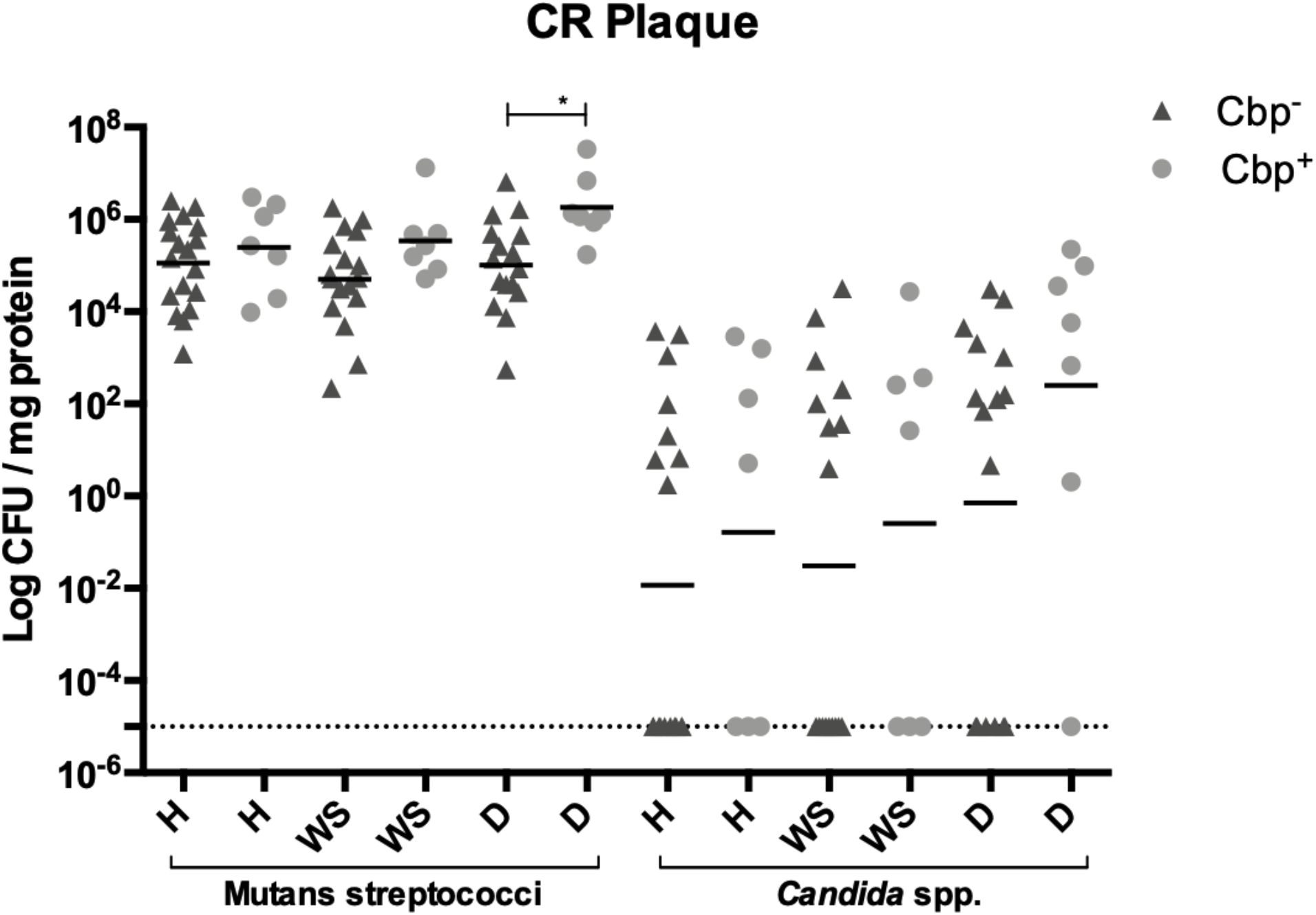
Microbiological analysis of plaque samples collected from subjects with active recurrent caries (CR group). Graph shows Mutans streptococci and *Candida* spp. counts in healthy plaque (H), white spot lesion plaque (WS) and cavitated dentin plaque (D) of CR subjects in relation to infection with *S. mutans* strains harboring either *cnm* or *cbm* genes (Cbp^+^). The CFU counts of the Cbp^*+*^ and Cbp^−^ CR subjects were compared using general linear mixed modeling. Data shown are mean ± SD values of log total colony count of Mutans streptococci and *Candida* spp. colonies.

### Salivary microbiome analysis revealed that *Streptococcus parasanguinis* is overrepresented in the CR group

We investigated the salivary microbiome using the Nanopore MinIon platform to gain additional insight on the microbiota associated with ECC recurrence and whether other biomarkers of caries recurrence could be found. After filtering out *H. sapiens* sequences, which consisted of about 87% of all obtained sequences, and samples with coverage under 70%, the final dataset comprised of 53,953 long-read sequences distributed in 28 samples (CF: n=10; CE-NR: n=8; CR: n=10) (Supplemental Table 2). The average number of sequences per sample was 1,926 and the median was 698 with a total of 703 bacterial species detected. No sequences belonging to fungi, viruses or archaea were detected at the coverage level obtained within this dataset. The alpha diversity mean of children with ECC recurrence was significantly greater as compared with caries free children (p=.011) but not with children in the CE-NR group (p=.57) (Fig. 3). The most abundant operational taxonomic units (OTUs) found in the three groups were *Streptococcus mitis* and *Streptococcus salivarius* (Fig. 4). Even though *S. mutans* was detected in higher abundance in CR subjects than in the other two groups, it comprised only a small proportion of the salivary microbiome (less than 5%) (Fig. 4). However, differential abundance analysis by the ALDEx2 ANOVA-like method revealed that *Streptococcus parasanguinis* was relatively more abundant in CR subjects than among the CF and CE-NR groups. The relative abundance of this microorganism was 6.53 and 4.28 in CR and CF subjects, respectively, with an effect of 0.62 and a significant difference between them (p=.05). When CR and CE-NR groups were compared, a marginal difference was observed in which *S. parasanguinis* relative abundance was 6.48 to CR and 4.72 to CE-NR with an effect of 0.58 (p=.06).

**Figure 3.**
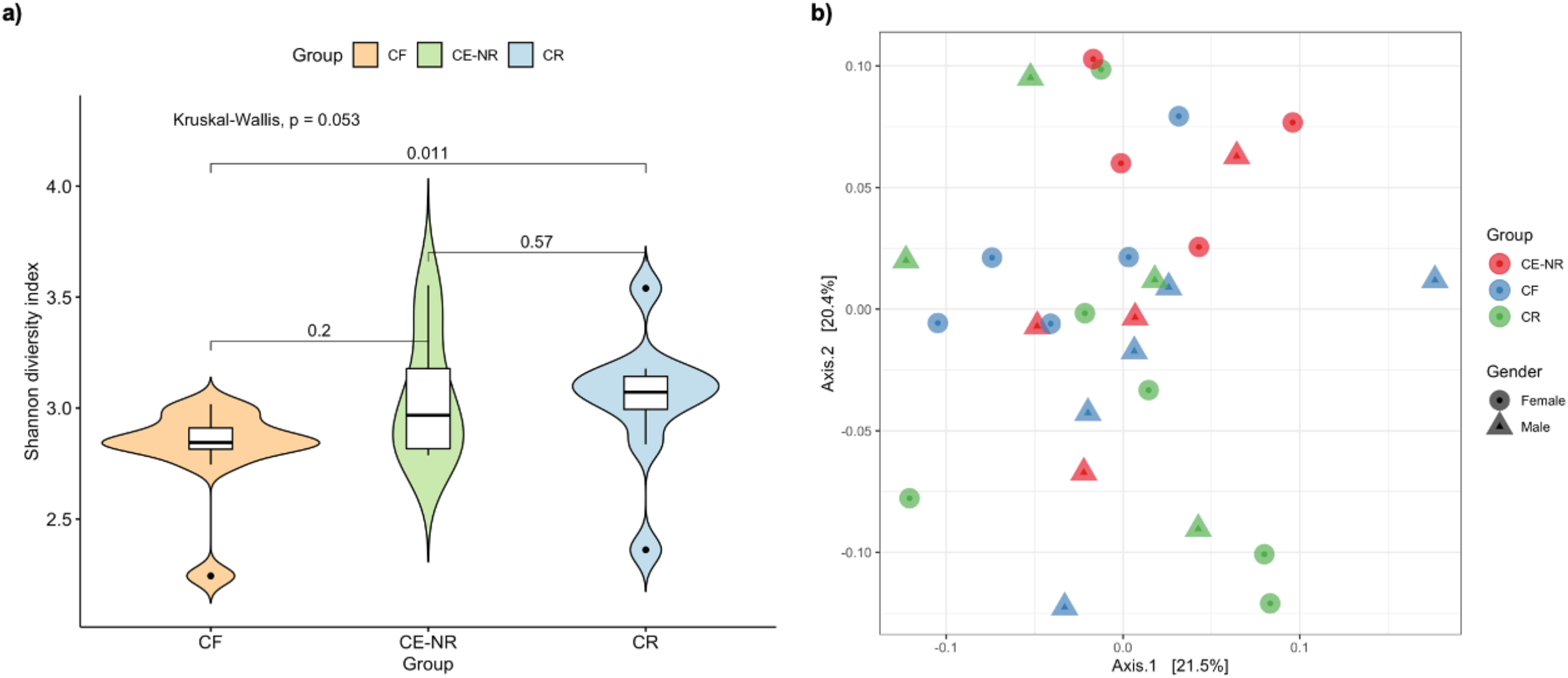
Shannon alpha diversity and Beta diversity metrics of the salivary microbiome of children who were caries free (CF), caries experienced – no recurrence (CE-NR) and had active recurrent caries (CR). (A) Violin plot depicting Shannon alpha diversity measures for each sample category was analyzed by paired Kruskal-Wallis test. CR group has a significantly higher alpha diversity when compared to CF and CE-NR. (B) Beta diversity was compared by using Principal Coordinate Analysis (PCoA) and the strength of the differences were tested by Permutational Analysis of Variance (PERMANOVA). No differences were observed in Beta diversity among the groups.

**Figure 4.**
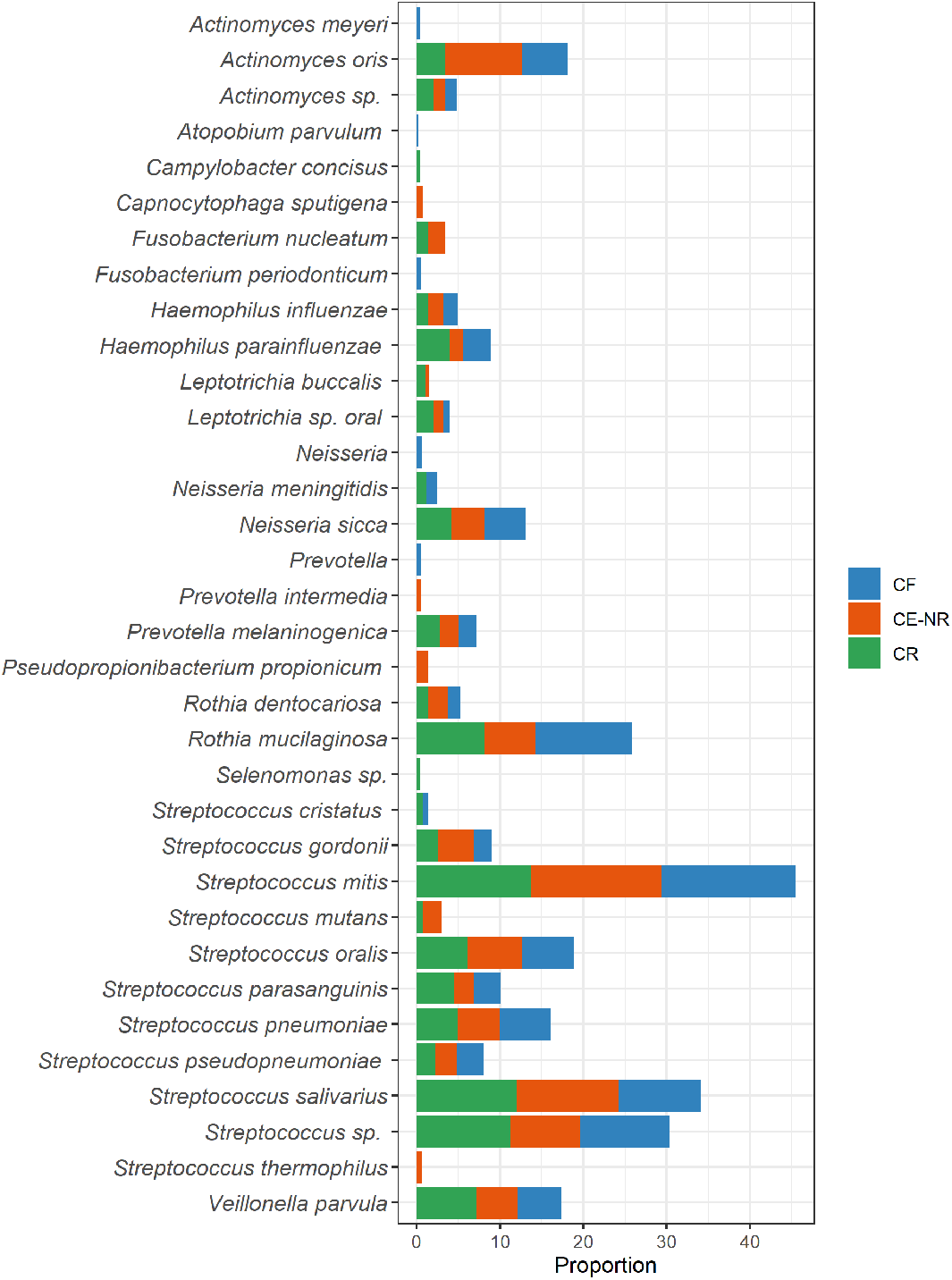
Distribution of the top 35 most abundant taxa across the caries free (CF), caries experienced - no recurrence (CE-NR) and active recurrent caries (CR) groups using rarefied taxon counts. The chart shows mean taxon counts for sample group. Means are shown as relative proportions.

The microbial communities from children of different caries statuses were compared by using multivariate PERMANOVA and Bray-Curtis distances, which showed no significant difference in beta diversity (R^2^=.097; p=.189) (Fig. 2). Age (*R*^2^=.047; *p*=.252), sex (*R*^2^=.018; F=.488□; *p*=.919) and caries status (*R*^2^=.078□; F Model=1.043; *p*-value=.919) had no significant association with oral bacterial composition.

## DISCUSSION

Despite increased surveillance and better treatment approaches^3^, ECC continues to be a significant health problem^2^ that has a high recurrence rate. While the association of *S. mutans* and *C. albicans* in caries initiation and progression is well established^19^, much less is known about the characteristics of the microbiota associated with ECC recurrence. Therefore, new predictors of ECC recurrence with high prognostic value are still needed^20^. In the present study, we observed an association of co-infection with *S. mutans,* especially with Cbp^+^ strains, and *C. albicans* with caries recurrence in children that received restorative treatment for ECC and developed new dentin caries lesions within 6 months post-treatment.

Previously, our group showed that the expression of Cbps confers *S. mutans* with the ability to bind more avidly to collagen-rich tooth surfaces such as dentin and roots, as well as to invade oral epithelial cell lines^11,21^, which also contributes to *S. mutans* niche expansion beyond the tooth enamel^22^. Here, Cbp^+^ *S. mutans* strains were only found in children with caries recurrence and classified as S-ECC because they were younger than 3 years old. Based on the ability of Cbp^+^ strains to colonize oral epithelial cells^11^, it is possible that oral infection by Cbp^+^ *S. mutans* in these children occurred before tooth eruption, which would have allowed *S. mutans* to colonize the dental biofilm immediately upon tooth eruption. In an animal model of caries, Miller *et al*.^11^ demonstrated that infection with *cnm*^+^ *S. mutans* is associated with increased *S. mutans* load in dental plaque and with caries severity, suggesting a role for Cnm in the assembly of the cariogenic biofilm^11^. Furthermore, clinical studies have demonstrated that infection with *cnm*^+^ *S. mutans* is associated with increased caries risk in children^9,10^. We speculate that because Cbp^+^ *S. mutans* can colonize multiple niches in the oral cavity, possibly including soft tissues, these strains can rapidly recolonize oral surfaces after oral hygiene, prophylaxis and intervention measures, ultimately resulting in recalcitrant infections.

In addition to *S. mutans*, the presence of *Candida* species in the oral cavity has been associated with dental caries, with *C. albicans* being the most prevalent species present in caries-active children^15,23,24^. In our cohort, CR subjects harbored highest levels of *Candida* spp. in plaque, especially in carious dentin plaque. Because collagen type I is the main organic component of dentin^25^ and *C. albicans* is highly aciduric and able to efficiently adhere to and degrade collagen, it is therefore not entirely unexpected that *C. albicans* is found in dentin carious lesions^26–28^. Also, *in vitro* studies showed that clinical isolates of *C. albicans* can promote tooth demineralization with consequent loss of hardness^29^, a finding that was supported by *in vivo* studies showing that rats infected with *C. albicans* developed carious lesions^18,30^. Notably, *S. mutans* and *C. albicans* have a synergistic interaction that promotes assembly of hypercariogenic biofilms resulting in the most aggressive forms of caries in children^14–17^ and in animals^18^. Moreover, *S. mutans* and *C. albicans* together are recalcitrant to surgical interventions (i.e. extensive dental treatment usually provided in hospital operating rooms) followed by prophylactic chlorhexidine treatments^17^ thereby contributing to increased risk of caries recurrence. In addition to synergistic interactions that promote biofilm accumulation^18^, *C. albicans* possesses several traits making it a desirable neighbor for *S. mutans*. More specifically, *C. albicans* utilizes lactic acid, a byproduct of *S. mutans* metabolism, as energy source, is highly acidogenic and aciduric, and reduces oxygen tension of the biofilm^31^. At the same time, *S. mutans* secretes glucosyltransferase B that binds to the surface of *C. albicans* making it a *de facto* glucan producer when sucrose is present, contributing to accumulation of biofilm matrix^32^. Strikingly, we observed that CR subjects infected with Cbp^+^ *S. mutans* had at least 13-fold more *C. albicans* and 8-fold more Mutans streptococci counts in dentin than subjects infected with Cbp^−^ *S. mutans*. Thus, it seems that co-infection with Cbp^+^ *S. mutans* and *C. albicans* results in even more potent synergistic cross-kingdom interactions. Studies investigating the contribution of Cbps to the establishment of *S. mutans* and *C. albicans* cariogenic biofilms are currently underway.

In accordance with previous studies^7,33,34^, our data demonstrated that serotype *c* is the most prevalent *S. mutans* serotype found in our cohort. Although most subjects of this study were infected with a single *S. mutans* serotype, co-infection with serotypes *e* and *k* was associated with caries recurrence, which is in alignment with studies showing that infection by more than one *S. mutans* serotype is associated to increased caries risk^34^. Even though Cbp^+^ strains have been commonly found in the less common serotypes *e, f* and *k*^10,35^, the majority of the Cbp^+^ *S. mutans* isolates identified in our study belonged to serotype *c*^7^. The differences observed in serotype prevalence of Cbp^+^ strains between previous studies and ours may relate to how little is known about the epidemiology of Cbp^+^ *S. mutans* in the Americas, as most previous studies were done with European and Asian cohorts^9,10,36^.

Using the Nanopore MinIon platform, we showed that the salivary microbiome of the CR group has greater alpha diversity than the CF group. This finding was unexpected as previous publications showed that in greater caries incidence, there is a reduction in diversity and an enrichment for aciduric and acidogenic species^37–39^. It should be noted that, to our knowledge, this is the first time that the Nanopore sequencing platform was used to study oral microbiome of caries recurrence. One possibility for this unexpected finding is that, when compared to the other sequencing platforms used in previous oral microbiome studies, Nanopore sequencing generates less bias as it does not require enrichment and 16S amplification steps. Moreover, the Nanopore platform generates long reads allowing appropriate species-level assignments whereas for 16S-based sequencing platforms, species assignment is not always possible and often times bacterial identifications are made at genus level.

Similar to previous a microbiome studies^40,41^, our salivary microbiome data could not identify specific taxa that could serve as biomarkers of health and disease. However, we found that *S. parasanguinis* was relatively more abundant in CR subjects than in the CF and CE-NR groups. Interestingly, studies comparing dental plaque from caries-active and caries-free subjects also observed that *S. parasanguinis* was over-represented in caries-active patients^37,42^. Future studies are warranted to address the role of *S. parasanguinis* in caries^37^.

Collectively, this study indicates that children colonized with Cbp^+^ *S. mutans* may be predisposed to recalcitrant caries infections, and that oral co-infection with Cbp^+^ *S. mutans* and *C. albicans* might be even harder to eradicate than *S. mutans* alone or Cbp^−^ *S. mutans* and *C. albicans*. The concept that ECC recurrence could be attributed to persistent co-infection with Cbp^+^ *S. mutans* and *C. albicans* is novel and warrants further investigation. If this relationship is confirmed, a paradigm shift can be made towards ECC prevention and treatment through the integration of this information into the assessment of caries risk. For example, simple tests involving oral swabs can be used to screen infants and toddlers for the presence of Cbp^+^ *S. mutans* and *Candida* species. Then, early interventions could be adopted to specifically target these microorganisms in children with ECC given the recalcitrant nature of this infection in this population.

## METHODS

### Recruitment and Enrollment

The human subjects protocol was reviewed and approved by the Institutional Review Board of the University of Florida (IRB201600851). This cross-sectional study conforms to STROBE guidelines (Strengthening the Reporting of Observational Studies in Epidemiology) for cohort studies. Children ranging from 6 to 72 months, in their primary dentition, were enrolled in the study. All subjects were patients at the University of Florida College of Dentistry (UFCD) Department of Pediatric Dentistry. Written informed consent was obtained from at least one parent or legal guardian of the subjects before collection of the samples. Subjects that were in good general health and had not used antibiotics at least 1 month before their appointment were selected based on their overall baseline caries experience. Other exclusion criteria included: any medical conditions requiring pre-medication prior to dental treatment, immunosuppression or any type of compromised immune system, and orthodontic or intra-oral orthopedic appliances. These subjects were then categorized into one of three groups based on the American Academy of Pediatric Dentistry (AAPD) criteria: (i) Caries Free group (CF; no visible caries), (ii) Caries Experienced - No Recurrence group (CE-NR; prior ECC treatment with no visible caries 6-months post treatment), and (iii) Caries Active - Recurrence group (CR; prior ECC treatment with visible dentin caries lesions within 3 to 6-months post treatment). Using AAPD criteria, subjects from the groups CE-NR and CR were also classified regarding ECC severity.

### Specimen Collection

All enrolled subjects donated saliva and plaque samples from healthy enamel surfaces (H). For CR patients, additional plaque samples from a white spot lesion (WS) on the edge of a carious lesion and from a cavitated lesion that visibly extends into the dentin (D) were collected. Approximately 1 ml of unstimulated saliva was obtained through a pediatric mucus trap attached to a vacuum pump^43^. All plaque samples were collected using a sterile dental micro-applicator brush, which was then deposited into tube containing 1 ml sterile 0.1M glycine buffer (pH 7) solution.

### Microbiological Analysis

Saliva and plaque samples were serially diluted in 1 ml of 0.1M glycine buffer solution and plated on mitis-salivarius agar (Becton, Dickinson and Company BBL^™^, Franklin Lakes, NJ, USA) supplemented with 0.388 mg/ml bacitracin (MSB) for quantification and isolation of Mutans streptococci^44,45^. Saliva samples were also plated onto tryptic soy agar + 5% sheep blood agar (TSA II; Moltox, Boone, NC, USA) for quantification of total bacteria. MSB and TSA II plates were incubated at 37°C in a 5% CO_2_ atmosphere for 48 hours. The TSA II plates were transferred to an aerobic incubator for an additional 24 hours to allow for growth of aerobes. Eight *S. mutans*-like colonies from each sample (saliva, H, WS and D) were picked from the MSB plates and patched onto tryptic soy agar (Becton, Dickinson and Company BBL^™^, Franklin Lakes, NJ, USA) plates for further analysis. Salivary counts of Mutans streptococci were normalized to total bacterial counts. Also, plaque counts of Mutans streptococci were normalized to total plaque protein content determined by bicinchoninic acid (BCA) protein assays. The identity of isolated *S. mutans*-like colonies was determined by PCR using *S. mutans*-specific primers^46^. Then, for the confirmed *S. mutans* colonies, the serotype and the presence of genes coding for the collagen and laminin binding proteins, *cnm* and *cbm,* were determined by PCR using serotype-specific primers^33,34,47^ and *cnm* and *cbm*-specific primers^35,48^ (Supplementary Table S1). UA159 (serotype *c*; *cnm*^-^), B14 (serotype *e; cnm*^+^), OMZ175 (serotype *f*; *cnm*^+^), and OM88X (serotype *k*; *cbm*^+^) were included as controls in the PCR reactions.

The prevalence of *Candida* spp. was determined by plating serial dilutions of saliva and plaque samples onto Sabouraud agar (Becton, Dickinson and Company BBL^™^, Franklin Lakes, NJ, USA) supplemented with 0.4 mg/ml chloramphenicol^15^. Salivary counts of *Candida spp* were normalized to total bacterial counts. Also, plaque counts of *Candida spp* were normalized to total plaque protein content determined by bicinchoninic acid (BCA) protein assays. Then, eight *Candida* spp. colonies from each sample (saliva, H, WS and D) were streaked onto HardyCHROM^™^ agar (Hardy Diagnostics, Santa Maria, CA, USA) for presumptive identification of the four main human-associated *Candida* species (*C. albicans, C. tropicalis, C. glabrata, and C. krusei*).

### Salivary Microbiome Analysis

The salivary microbiome was investigated using the Nanopore MinION sequencing platform (Oxford Nanopore Technologies, Oxford, UK). Salivary DNA from each sample was extracted using the Quick-DNA^™^ Fungal/Bacterial Miniprep Kit following the manufacturer’s protocol (Zymo Research Corp, Irvine, CA, USA). Library preparation was performed using 400 ng of DNA from each sample and Rapid Barcoding Sequencing (SQK-RKB004, Oxford Nanopore Technologies, Oxford, UK) following manufacturer’s guidelines. Samples were pooled, cleaned and concentrated using AxyPrep FragmentSelect (Axygen, Union City, CA, USA). Next, 1□µl of rapid adapter was added to 10□µl of the pooled eluate and the library kept on ice until loaded onto the flow cell. Groups of 12 saliva samples were pooled to create one library. Shotgun metagenomic sequencing was carried out on a FLO-MIN 106D R9 MinION sequencer version. The Barcoding and WIMP workflows were employed using default Q-score ≥7. Each MinION cell was run for 48□ hours. The result files from the 3 workflows (12 patients in each workflow) were combined in order to compare the microbiota of all samples. The Raw sequences were deposited in the Sequence Read Archive (SRA) BioProject (accession number PRJNA679743).

### Statistical Analysis

The counts of Mutans streptococci and *Candida* spp. in the plaque and saliva among the three caries status groups were compared using general linear mixed modeling. To improve the normal distribution of the concentrations for regression modeling while also accounting for the presence of values of zero among the plaque and saliva assays, log transformations that included a constant in the form of, for example, log Mutans streptococci in plaque = log (Mutans streptococci counts in plaque + 0.000001) were used. Because there were age differences among the three groups, age in months was included as a covariate in the mixed models. Progression analyses were conducted using the MIXED procedure in the SAS statistical analysis package, version 9.4 (SAS Institute, Cary, NC, USA).

Microbial identification was carried out using WIMP analysis tool. The resulting table with the classification of each long Nanopore read was imported into the R environment. A contingency table was built and used for downstream analysis. The dataset was filtered by removing *Homo sapiens* DNA sequences and only samples with sequence coverage greater than 70% were kept. After rarefying the samples to the same number of sequences, Shannon diversity index estimator were calculated using the “phyloseq” package, and then plotted using the “ggpubr” package, both in the R environment. The alpha diversity measurements were tested for normality using the Shapiro-Wilk test, and differences among the categories were determined with the Kruskall-Wallis test.

Differences in microbial community structure were visualized by Principal Coordinate Analysis (PCoA) and the strength of the differences tested by Permutational Analysis of Variance (PERMANOVA). Briefly, the centered log ratio transformation was applied and the transformed dataset used to construct a dissimilarity matrix generated by euclidean distance. The matrix was ordinate by Principal Coordinate Analysis (PCoA) and the adonis function was used to calculate the Permutational Multivariate Analysis of Variance (PERMANOVA) with the vegan package. The differential abundance analysis among groups was performed using ALDEx2^49^.

## Data Availability

All strains are available upon request. The Raw sequences were deposited in the Sequence Read Archive (SRA) BioProject (accession number PRJNA679743).

## Acknowledgements

We thank the pediatric dentistry residents Drs. Joshua Cline, Latoya Jones, Nikki Darbani, Max Rudie, Andrés Alvarez, Kiersten Pannel and Catherine Graham for excellent clinical assistance in collecting patient samples. We thank Dr. Jessica Kajfasz for critically revising the text. This work was supported by DE022559, 5R90DE022530-09 and Colgate-Palmolive CARE award.

## Author contributions

B.A.G. and N.C.A., contributed to data acquisition, analysis, and interpretation; and drafted and critically revised the manuscript. S.L.T. and L.F.W.R., contributed to data analysis and interpretation; and critically revised the manuscript. L.C.F.M., contributed to conception, design, and critically revised the manuscript; J.A.L., contributed to conception and critically revised the manuscript; J.A., contributed to conception, design, data analysis and interpretation and critically revised the manuscript. All authors gave final approval and agree to be accountable for all aspects of the work.

