## supplemental tables for "Association of *Streptococcus mutans* harboring bona-fide collagen binding proteins and *Candida albicans* with early childhood caries recurrence"

| Supplementary table S1. PCR primers used for *S. mutans* detection, serotype determination, and *cnm*/*cbm* detection | | | | |
| --- | --- | --- | --- | --- |
| **Primer** | **Purpose** | **Sequence (5'-3')** | **Expected size (bp)** | **Reference** |
| sm479F | *S. mutans* detection | TCG CGA AAA AGA TAA ACA AAC A | 479 | 46 |
| sm479R |  | gcc cct tca cag ttg gtt ag |  |  |
| SerotypeC-F | Serotype *c* determination | CGG AGT GCT TTT TAC AAG TGC TGG | 727 | 33, 34 |
| SerotypeC-R |  | AAG CAC GGC CAG CAA ACC CTT TAT |  |  |
| SerotypeE-F | Serotype *e* determination | CCT GCT TTT CAA GTA CCT TTC GCC | 517 | 33, 34 |
| SerotypeE-R |  | CTG CTT GCC AAG CCC TAC TAG AAA |  |  |
| SerotypeF-F | Serotype *f* determination | CCC ACA ATT GGC TTC AAG AGG AGA | 316 | 33, 34 |
| SerotypeF-R |  | TGC GAA ACC ATA AGC ATA GCG AGG |  |  |
| CEFK-F | Serotype *k* determination | ATT CCC GCC GTT GGA CCA TTC C | 294 | 33, 47 |
| K-R |  | CCA ATG TGA TTC ATC CCA TCA C |  |  |
| cnm541-F | Identification of *cnm* | AGC TGA GGT TAC TGT CGT TA | 361 | 35 |
| cnm901-R |  | CAG GAT TGT CAA CTT TAG TC |  |  |
| cbm-EF | Identification of *cbm* | AGC TGA AGT TAG TTA AAA CCT GCT TC | 393 | 48 |
| cbm-ER |  | TAG GAT CAT CAA CGT CAA GTA CAC GA |  |  |

| Supplementary table S2. Characteristics of the sequenced samples | | | | | | |
| --- | --- | --- | --- | --- | --- | --- |
| Sample | Group | Biological sex | Age | Total Reads Homo sapiens | Total Reads Bacteria | Coverage |
| p78 | CR | Female | 26 | 29612 | 10351 | 0.96 |
| p11 | CF | Female | 26 | 1825 | 639 | 0.93 |
| p19 | CR | Male | 35 | 10736 | 4466 | 0.98 |
| p2 | CR | Female | 20 | 569 | 111 | 0.87 |
| p24 | CF | Male | 36 | 7577 | 4625 | 0.99 |
| p32 | CE-NR | Male | 29 | 168 | 59 | 0.71 |
| p34 | CF | Male | 32 | 301 | 343 | 0.91 |
| p36 | CF | Female | 30 | 2578 | 475 | 0.91 |
| p38 | CR | Female | 26 | 8582 | 714 | 0.94 |
| p39 | CF | Female | 30 | 413 | 82 | 0.84 |
| p45 | CF | Male | 18 | 868 | 235 | 0.88 |
| p47 | CR | Male | 30 | 3531 | 929 | 0.96 |
| p51 | CR | Female | 23 | 12864 | 683 | 0.92 |
| p54 | CE-NR | Female | 35 | 16320 | 3570 | 0.98 |
| p59 | CF | Female | 26 | 6556 | 548 | 0.92 |
| p6 | CR | Male | 36 | 6634 | 3308 | 0.97 |
| p62 | CE-NR | Male | 57 | 2604 | 298 | 0.85 |
| p65 | CR | Male | 48 | 163304 | 10596 | 0.99 |
| p66 | CE-NR | Male | 41 | 11689 | 1138 | 0.97 |
| p67 | CE-NR | Female | 59 | 5462 | 1324 | 0.96 |
| p68 | CE-NR | Female | 36 | 7255 | 1568 | 0.96 |
| p7 | CF | Male | 34 | 838 | 407 | 0.92 |
| p79 | CE-NR | Male | 48 | 779 | 154 | 0.88 |
| p88 | CE-NR | Female | 41 | 10064 | 719 | 0.93 |
| p95 | CR | Female | 57 | 40112 | 4034 | 0.98 |
| p96 | CR | Female | 40 | 3083 | 315 | 0.91 |
| p97 | CF | Male | 55 | 3173 | 154 | 0.81 |
| p118 | CF | Female | 56 | 3362 | 2108 | 0.97 |
